# Early prediction of high risk gestational diabetes mellitus via machine learning models

**DOI:** 10.1101/2020.03.26.20040196

**Authors:** Yan-Ting Wu, Chen-Jie Zhang, Ben Willem Mol, Cheng Li, Lei Chen, Yu Wang, Jian-Zhong Sheng, Jian-Xia Fan, Yi Shi, He-Feng Huang

**Author notes:** Yan-Ting Wu and Chen-Jie Zhang contributed equally to this work. Co-corresponding author: He-Feng Huang, International Peace Maternity and Child Health Hospital, School of Medicine, Shanghai Jiao Tong University, No.910, Hengshan Road, Shanghai, 200030, China.cn, Yi Shi, Key Laboratory of Systems Biomedicine (Ministry of Education), Shanghai Center for Systems Biomedicine, Shanghai Jiao Tong University, Shanghai, China.

## Abstract

**Aims:** Gestational diabetes mellitus (GDM) is a pregnancy-specific disorder that can usually be diagnosed after 24 gestational weeks. So far, there is no accurate method to predict GDM in early pregnancy.

**Methods:** We collected data extracted from the hospital’s electronic medical record system included 73 features in the first trimester. We also recorded the occurrence of GDM, diagnosed at 24-28 weeks of pregnancy. We conducted a feature selection method to select a panel of most discriminative features. We then developed advanced machine learning models, using Deep Neural Network (DNN), Support Vector Machine (SVM), K-Nearest Neighboring (KNN), and Logistic Regression (LR), based on these features.

**Results:** We studied 16,819 women (2,696 GDM) and 14,992 women (1,837 GDM) for the training and validation group. DNN, SVM, KNN, and LR models based on the 73-feature set demonstrated the best discriminative power with corresponding area under the curve (AUC) values of 0.92 (95%CI 0.91, 0.93), 0.82 (95%CI 0.81, 0.83), 0.63 (95%CI 0.62, 0.64), and 0.85 (95%CI 0.84, 0.85), respectively. The 7-feature (selected from the 73-feature set) DNN, SVM, KNN, and LR models had the best discriminative power with corresponding AUCs of 0.84 (95%CI 0.83, 0.84), 0.69 (95%CI 0.68, 0.70), 0.68 (95%CI 0.67, 0.69), and 0.84 (95% CI 0.83, 0.85), respectively. The 7-feature LR model had the best Hosmer-Lemeshow test outcome. Notably, the AUCs of the existing prediction models did not exceed 0.75.

**Conclusions:** Our feature selection and machine learning models showed superior predictive power in early GDM detection than previous methods; these improved models will better serve clinical practices in preventing GDM.

**Research in Context section:** *Evidence before this study:*

1. A hysteretic diagnosis of GDM in the 3rd trimester is too late to prevent exposure of the embryos or fetuses to an intrauterine hyperglycemia environment during early pregnancy.
2. Prediction models for gestational diabetes are not uncommon in previous literature reports, but laboratory indicators are rarely involved in predictive indicators.
3. The penetration of AI into the medical field makes us want to introduce it into GDM predictive models.

*What is the key question?:* Whether the GDM prediction model established by machine learning has the ability to surpass the traditional LR model?

*Added value of this study:*

1. Using machine learning to select features is an effective method.
2. DNN prediction model have effective discrimination power for predicting GDM in early pregnancy, but it cannot completely replace LR. KNN and SVM are even worse than LR in this study.

*Implications of all the available evidence:* The biggest significance of our research is not only to build a prediction model that surpasses previous ones, but also to demonstrate the advantages and disadvantages of different machine learning methods through a practical case.

## Introduction

Gestational diabetes mellitus (GDM) is a common complication during pregnancy, which is very different from pure overt diabetes [1], and it affects up to 15% of pregnancies in the world [2]. Hyperglycemia is not, by itself, life-threatening for pregnant women, but it can be harmful to the fetus. Fetal symptoms include polyhydramnios, stillbirth, premature delivery, macrosomia, fetal hyperinsulinemia, and clinical neonatal hypoglycemia. The American Diabetes Association (ADA) and International Association of Diabetes and Pregnancy Study Groups (IADPSG), recommends diagnosing GDM via 2h, 75g oral glucose tolerance test (OGTT) at 24-28 weeks of pregnancy [3, 4]. However, a hysteretic diagnosis of GDM in the 3^rd^ trimester is too late to prevent exposure of the embryos or fetuses to an intrauterine hyperglycemia environment during early pregnancy. There is accumulating evidence indicating that exposure of embryos or fetuses to a hyperglycemic environment in the uterus can lead to chronic health problems later in life [5], including obesity, diabetes, and cardiovascular diseases [6-8]. Our previous studies indicate that insulin therapy after GDM diagnosis cannot fully protect offspring from diet-associated metabolic disorders in adulthood [9]. Therefore, it is essential to establish a prediction model to forecast the possibility of GDM during early pregnancy and to provide lifestyle interventions prior to diagnosis in the 3^rd^ trimester.

Previous research has sought to find a threshold value of fasting plasma glucose (FPG) in the first trimester through large sample studies. While elevating diagnostic criteria from FPG ≥5.1 mmol/l to FPG ≥6.1 mmol/l can obtain nearly 100% specificity, the relatively low sensitivity (1%) greatly limits the feasibility [10]. In recent years, some novel biomarkers have been reported as potential prediction parameters of GDM, including angiopoietin-like protein 8, plasma fatty acid-binding protein 4 and various adipokines [11-13]. However, these biomarkers cannot be easily obtained by hospitals undermining the prediction accuracies.

Exploration of prediction models based on multiple common risk factors, such as advanced maternal age, body mass index (BMI), and family history of diabetes, provides a new perspective on solving the problem [14]. There are still only a few studies, however, that combine demographic characteristics and laboratory testing into a prediction model. Recently, AI technology, especially supervised machine learning methods, were reported to demonstrate a powerful self-learning ability and improve accuracies in diabetic retinopathy and pediatric disease prediction/diagnosis [15]. In order to explore whether combining AI with prior clinical knowledge will boost the early prediction and diagnosis of diseases such as GDM, we developed feature selection-based machine learning models to predict the occurrence of GDM based on clinical features in the 1^st^ trimester.

## Methods

### Data source

We collected the 2017 obstetrical electronic medical record data from the International Peace Maternal and Child Health Hospital (IPH), Shanghai Jiao Tong University School of Medicine, which served as the training group. Women with pregestational diabetes mellitus and women who terminated their pregnancy before the 75g OGTT test (24-28 weeks of pregnancy) were excluded. To improve data quality, we further filtered samples and features/biomarkers whose missing value rate was >20%.

Candidate features containing sociodemographic characteristics, clinical features, and laboratory indexes in the first trimester were collected. We then collected and curated the 2018 obstetrical electronic medical record data, which served as the validation group to evaluate the prediction models. The GDM diagnostic criteria follow the IADPSG guidelines, i.e., FPG ≥ 5.1 mmol/L or 1 h ≥ 10 mmol/L or 2 h ≥ 8.5 mmol/L should be diagnosed as GDM. This study was approved by the Medical Ethical Committee of International Peace Maternity and Child Health Hospital, School of Medicine, Shanghai Jiao Tong University (No. GKLW2019-05).

### Feature selection by AI

To improve the downstream prediction power and provide a more cost-effective approach for clinical practice, the feature selection procedure was conducted to select a panel of biomarkers with the most discriminative power in distinguishing the GDM group from the control group. Specifically, we found that many features are highly correlated to each other, forming small clusters, as shown in Fig 1b, c, and Fig 2b, c; this means that a representative small cluster of features may provide sufficient discriminative power to enable early intervention. Including all redundant features from a cluster may undermine prediction effectiveness and efficiency. We followed the strategy that both our group and Xiao *et al*. previously demonstrated effective in gene selection [16, 17]. In detail, the feature selection is embedded in a cross-validation framework where both n-fold and leave-one-out cross-validation schemes are adopted. To test the performance of the n-fold cross-validation, given a feature selection method, the dataset is randomly partitioned into n equal parts where n-1 parts and the last part are treated as training dataset and testing dataset, respectively. The procedure is repeated n times so that each portion is evaluated as the testing dataset. The average classification accuracy over the whole process is considered to be the n-fold cross-validation classification accuracy of the feature selection method combined with the classifier. In this work, we chose n=10 after testing n=5, 10, 15, and 20, which all yielded similar outcomes. The random partition process was repeated 100 times. In total, the n-fold cross-validation classification accuracy is the average over a total of 1000 tests. For the leave-one-out cross-validation, each single data point (one sample) is held as a testing data point while the remaining data points are treated as the training dataset; the process is repeated for each data point and the classification accuracy is the average over the whole process. We adopted two classifiers in our study, the RBF kernel Support Vector Machine (SVM) classifier [18, 19] and the K-Nearest Neighbor (KNN) classifier [20].

**Fig 1.**
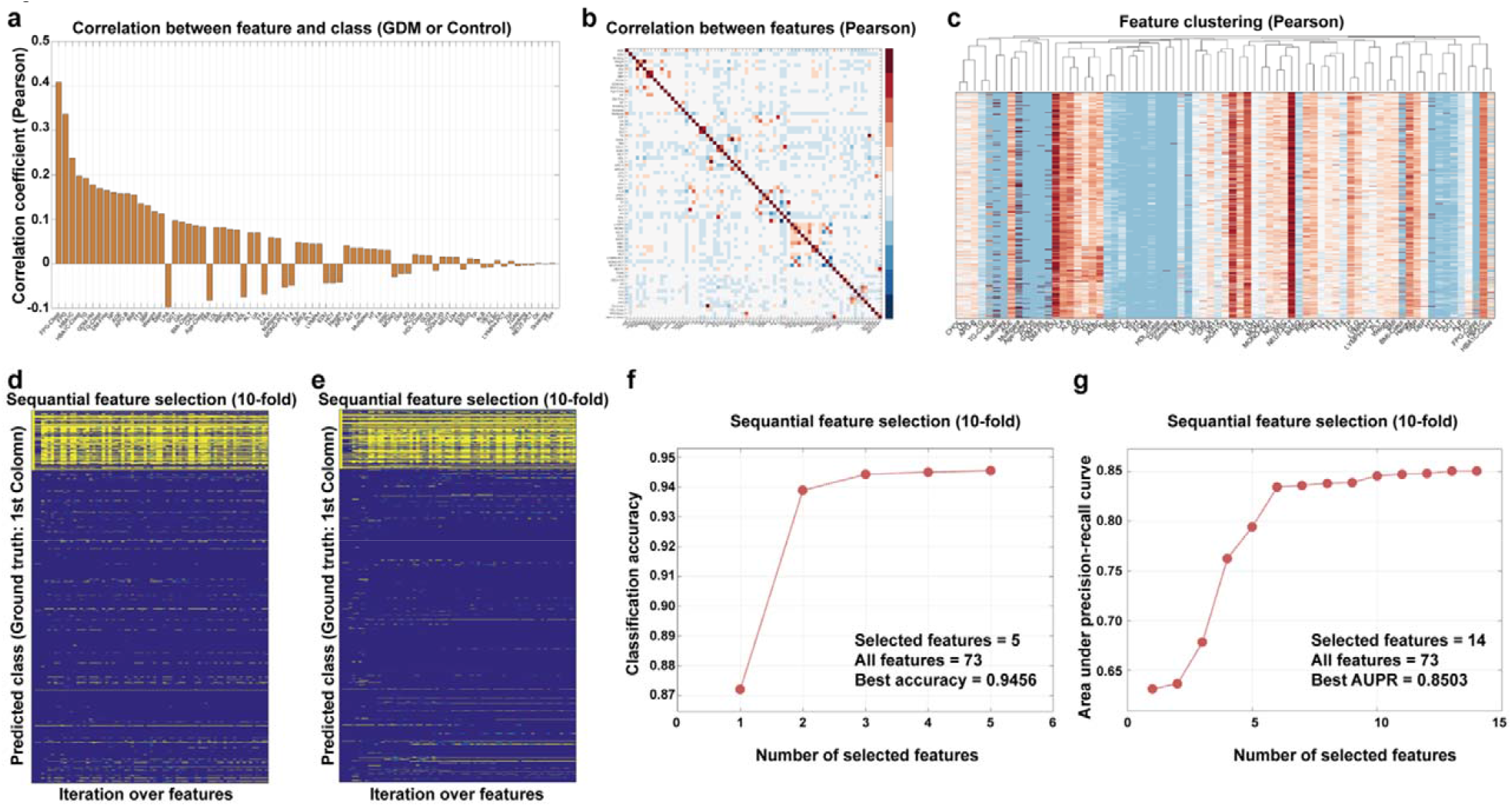
Feature selection results. (a) The Pearson correlation coefficients between each feature and the GDM/non-GDM label vector, overall the samples. Bar plots from left to right are absolute values from high to low. (b) The Pearson correlation coefficients between all the features over vectors of all the samples. (c) The feature-way hierarchical clustering results using distance metric based on Pearson correlation coefficients. (d and e). The 10-fold cross-validation based detailed prediction outcomes of each feature selection iteration. Yellow elements are predicted GDM cases and blue elements are predicted non-GDM cases (d) is seeking best accuracy. (e) is seeking AUC. (f) and (g) Feature selection trajectory guided by classification accuracy and AUC respectively, under 10-fold cross-validation framework.

**Fig 2.**
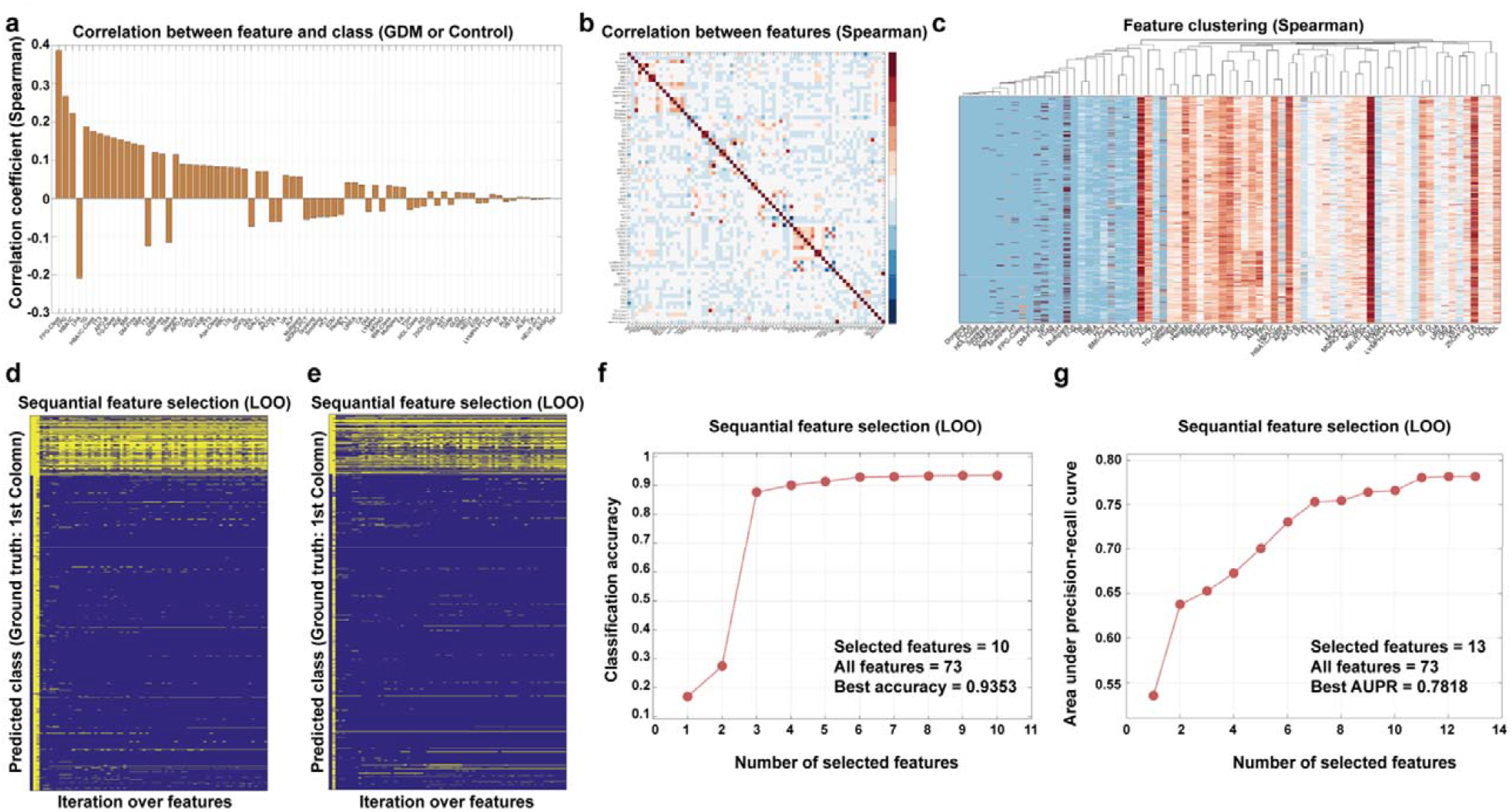
Feature selection results. (a) The Spearman correlation coefficients between each feature and the GDM/non-GDM label vector, over all the samples. Bar plots from left to right are absolute values from high to low. (b) The Spearman correlation coefficients between all the features over vectors of all the samples. (c) The feature-way hierarchical clustering results using distance metric based on Spearman correlation coefficients. (d) and (e) The leave-one-out cross-validation based detailed prediction outcomes of each feature selection iteration. Yellow elements are predicted GDM cases and blue elements are predicted non-GDM cases, (d) is seeking the best accuracy, (e) is seeking the best AUC. (f) and (g) Feature selection trajectory guided by classification accuracy and AUC respectively, under leave-one-out cross-validation framework.

All the features are initially sorted according to their absolute correlation coefficient (Pearson and Spearman) values with respect to the GDM/Control vector. The top-ranked feature is then included into the feature pool as the seed feature, and its discriminative power is determined by calculating the averaged prediction accuracy via the cross-validation process. Then, the second ranked feature is included into the feature pool and again a new prediction accuracy is calculated; if the new accuracy improves, the second ranked feature is included into the feature pool; otherwise, the third ranked feature will be tried. The process continues sequentially until all features have been considered. After this process, the features that are captured into the feature pool are treated as candidate features to be included in the final selected feature panel. Fig 1d, 1e and Fig 2d, 2e demonstrate the selected feature in each iteration. Fig 1f, 1g and Fig 2f, 2g plot the incremental trajectory of accuracies whenever including a contributing feature that stays in the selected feature pool.

### Prediction methods

Based on the selected feature panel, we tested four representative prediction methods: Logistic Regression (LR) [21], KNN [20], SVM [18,19], and Deep Neural Network (DNN) [22,23]. The effectiveness of each method is again tested using cross-validation. For the DNN classifier, a sequential model with two densely connected hidden layers is devised, followed by an output layer that returns a single, continuous value. For the hidden layers, we use a rectified linear unit as the activation function and RMS_prop_ as the optimizer. Mean squared error is used as a loss function while the mean absolute error is added as an evaluation metric. All-feature vectors and selected feature vectors are imported into the model separately and then trained for 1000 epochs.

The testing set mean abs error for 7-feature sets and all-feature sets are 0.17 and 0.12.

The update rule in RMS_prop_ is detailed as follows:

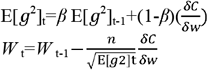

Where *E*(*g*^2^)denotes the moving average of squared gradients and ^δ*C*^/_δ*w*_ denotes the gradient of the cost function with respect to the weight. *n* is the□learning rate and *β* is the□moving average parameter.

For the LR classifier, during the training process, a sigmoid function of a given feature’s linear combination is trained. For the SVM classifier, we use an RBF (Gaussian) support vector model after considering linear kernel, polynomial kernel, and RBF kernels, where the parameters were set as default as used in the LIBSVM package. For the KNN classifier, we chose k=20 after testing k=1, 5, 10, 15, 20, 50, 100, so that the k-nearest neighbors’ majority voting is adopted as the prediction value.

### Model evaluation

All prediction models will draw ROC curves and calculate AUC to evaluate the discrimination of models. The Hosmer-Lemeshow (HL) test is used to evaluate the calibration of each model. Finally, the decision curve analysis (DCA) is introduced to evaluate the clinical application of each model.

## Results

### Sample size

In our training group, a total of 2,696 GDM patients were included in the case group and 14,123 pregnant women without GDM were included in the control group. 1,837 GDM patients and 13,155 pregnant women without GDM were included in the validation group.

### Features setting

The details of 73 alternative features containing sociodemographic characteristics, clinical features, and laboratory indexes in the first trimester are provided in Table 1, Table 2, and Table 3. Six features, including age, BMI, FPG, glycosylated hemoglobin A1C (HbA_1c)_, high density lipoprotein (HDL), and triglyceride (TG) were set as categorical variables apart from continuous variables.

**Table 1.**
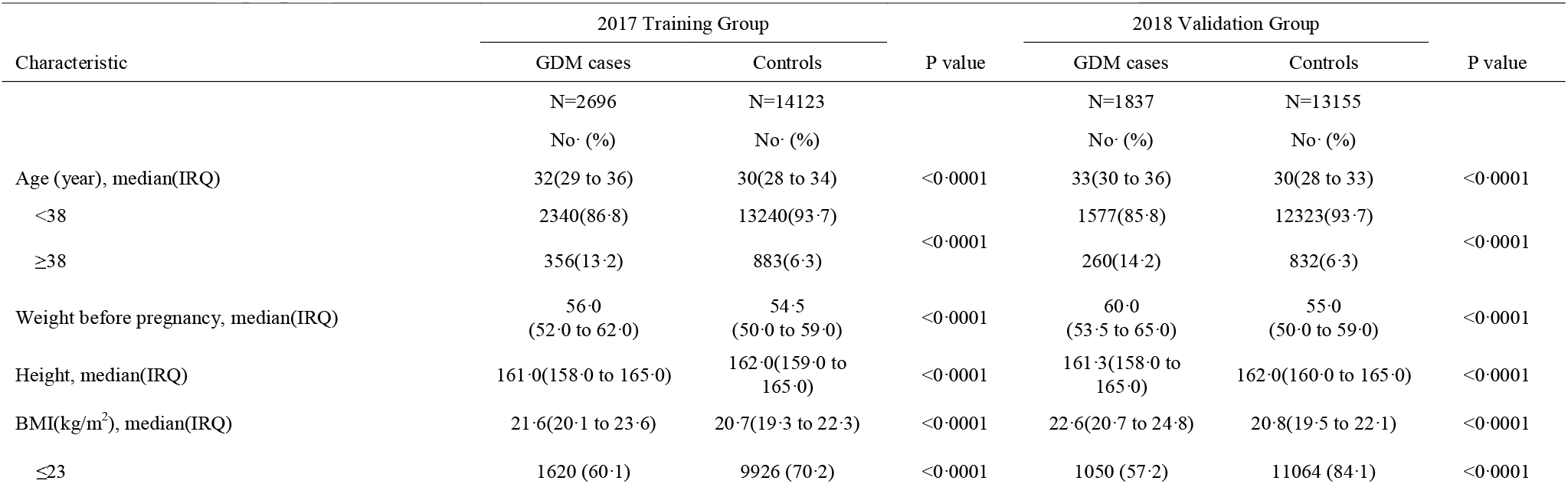
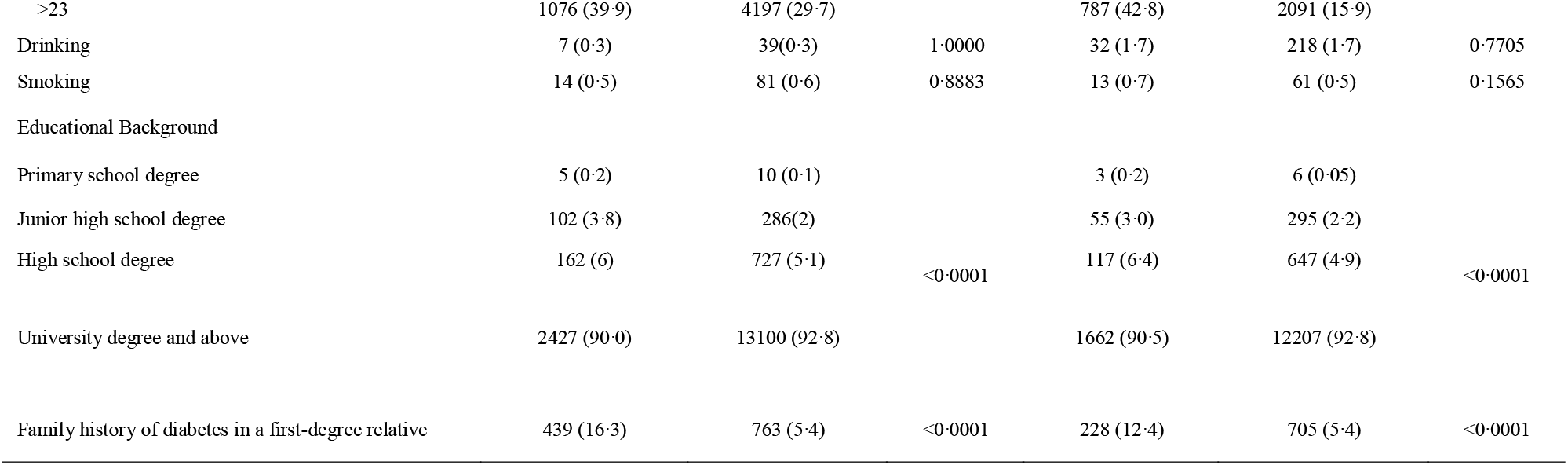
Sociodemographic characteristics of GDM and non-GDM cases.

**Table 2.** Clinical features of GDM and non-GDM cases in the first trimester.

| Characteristic | 2017 Training Group |  | P value | 2018 Validation Group |  | P value |
| --- | --- | --- | --- | --- | --- | --- |
|  | GDM cases | Controls |  | GDM cases | Controls |  |
|  | N=2696<br>No. (%) | N=14123<br>No. (%) |  | N=1837<br>No. (%) | N=13155<br>No. (%) |  |
| SBP (mmHg), median(IRQ) | 114(107 to 122) | 110(102 to 117) | <0.0001 | 116(107 to 124) | 110(102 to 117) | <0.0001 |
| DBP (mmHg), median(IRQ) | 71(65 to 77) | 68(62 to 73) | <0.0001 | 72(66 to 78) | 68(62 to 74) | <0.0001 |
| PCOS | 13 (0.5) | 30 (0.2) | 0.0195 | 29 (1.6) | 65 (0.5) | <0.0001 |
| Personal history of GDM | 132 (4.9) | 44 (0.3) | <0.0001 | 94 (5.1) | 44 (0.3) | <0.0001 |
| Natural pregnancy | 2180 (80.9) | 12324(87.3) | <0.0001 | 1432 (80.0) | 11462 (87.1) | <0.0001 |
| Multiple pregnancy | 110 (4.1) | 379(2.7) | <0.0001 | 77 (4.2) | 386 (2.9) | <0.0001 |
| Multipara | 1053 (39.1) | 4486 (31.8) | <0.0001 | 713 (38.8) | 4008 (30.5) | <0.0001 |

SBP, systolic blood pressure; DBP, diastolic blood pressure; PCOS, polycystic ovary syndrome

**Table 3.**
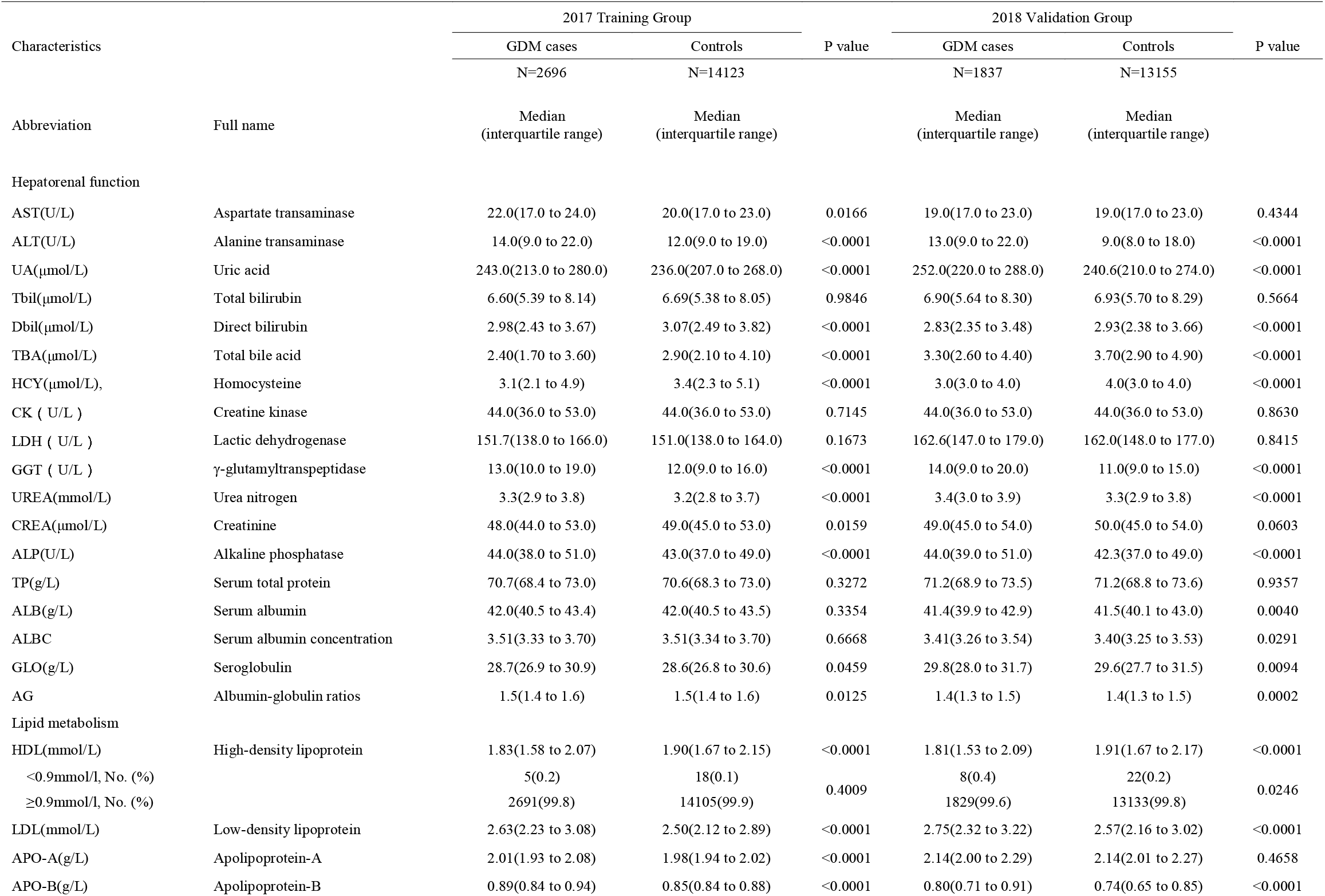
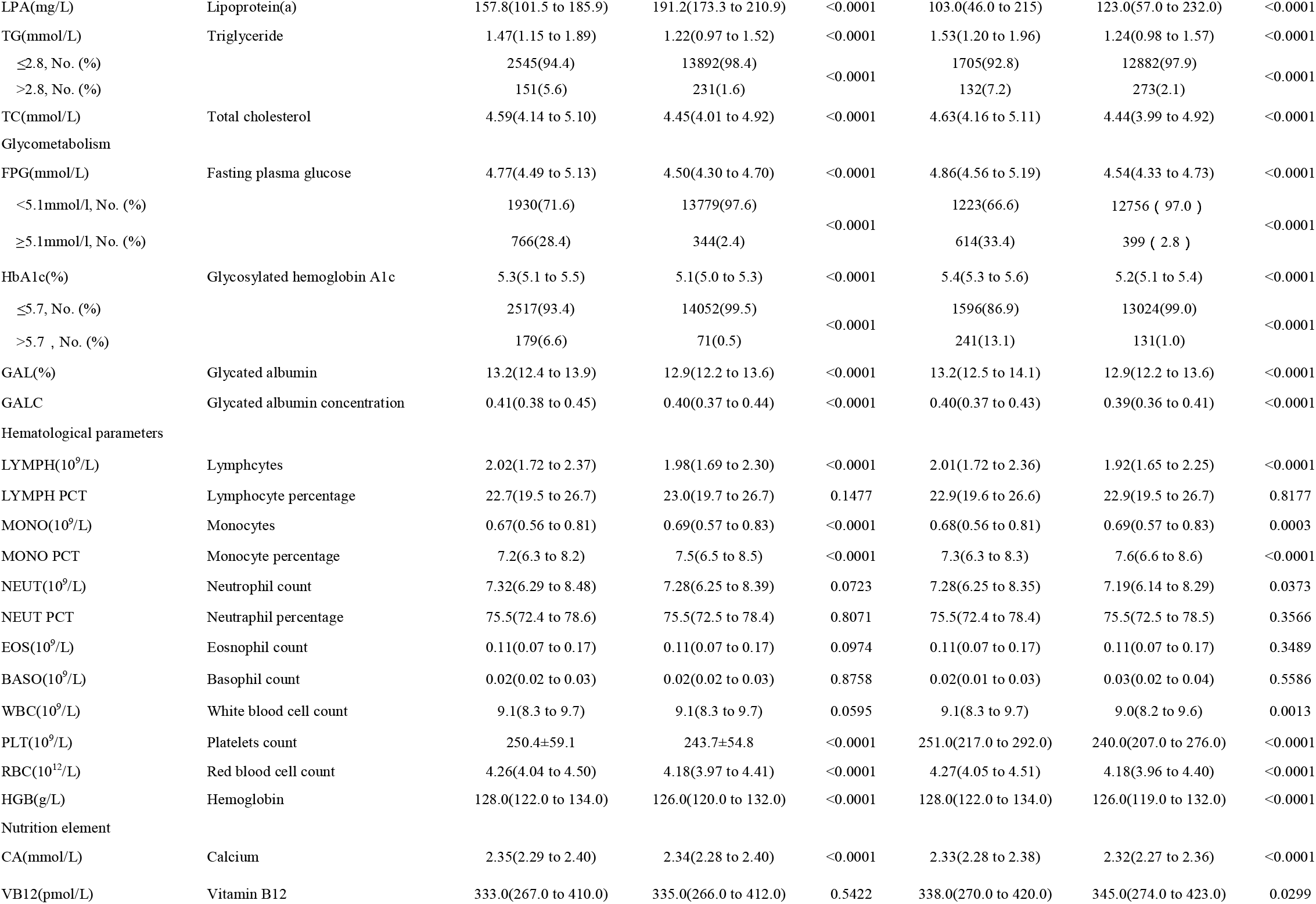
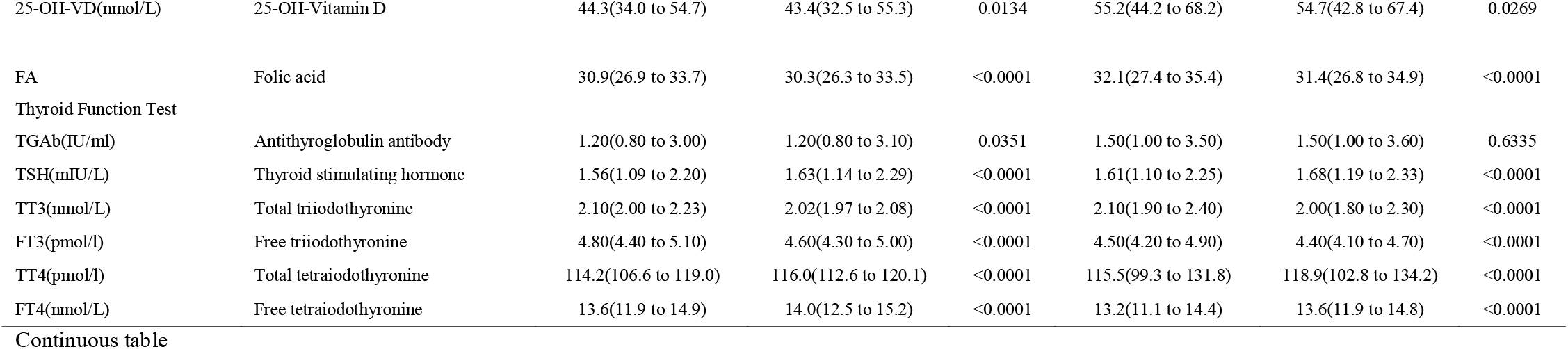
Laboratory indexes of pregnant women in early pregnancy.

### Feature selection

In the initial step of the feature selection procedure, all the features are sorted according to their absolute Pearson correlation coefficients and Spearman correlation coefficients with respect to the GDM/Control vector, as demonstrated in Fig 1a and Fig 2b. After the model-free sequential forward feature selection procedure, 17 features were captured in the feature pool, including age, age†, FPG, FPG†, HbA_1c_, HbA_1c_†, lipoprotein(a), apolipoprotein-A, apolipoprotein-B, TG, total triiodothyronine, total tetraiodothyronine, multiple pregnancy, multipara, smoking, family history of diabetes in a first-degree relative, previous GDM history (categorical variables denoted by †). More details are shown in Table 4. Based on prior clinical experience and a close examination of each feature, we further narrowed the feature set into a 7-feature panel to be used in further analyses and clinical practices. Interestingly, despite obesity being a well-known risk factor for GDM, AI did not choose BMI (a clinical indicator for obesity diagnosis), instead highlighting biochemical indicators that reflect the level of lipid metabolism, such as TG. This suggests that the use of biochemical indicators of lipid metabolism to predict GDM is more accurate than anthropometric indicators.

**Table 4.** Selecting features by KNN.

|  | 10-Fold<br>(Accuracy) | LOO<br>(Accuracy) | 10-Fold<br>(AUC) | LOO<br>(AUC) |
| --- | --- | --- | --- | --- |
| Selected<br>features | FPG† | Fasting plasma glucose | Fasting plasma glucose | FPG |
|  | Lipoprotein(a) | FPG† | FPG† | HbA1c |
|  | Total triiodothyronine | Lipoprotein(a) | Lipoprotein(a) | Family history of diabetes in a<br>first-degree relative |
|  | Age† | Total triiodothyronine | Total triiodothyronine | Triglyceride |
|  | Multiple pregnancy | Age | Triglyceride | Age |
|  |  | Total tetraiodothyronine | Age | Total triiodothyronine |
|  |  | APO-A | HbA1c† | Lipoprotein(a) |
|  |  | Multipara | Total tetraiodothyronine | Age† |
|  |  | Multiple pregnancy | Age† | Total tetraiodothyronine |
|  |  |  | APO-B | Multipara |
|  |  |  | Multipara | APO-A |
|  |  |  | Previous GDM | Multiple pregnancy |
|  |  |  | Multiple pregnancy | Previous GDM |
|  |  |  | Smoking |  |

- .12 †Categorical variable Age <38: 0, age≥38: 1; fasting plasma glucose<5.1 mmol/l: 0, FPG≥ 5.1 mmol/l: 1; - .13 HbA1c<5.7: 0, HbA1c ≥5.7: 1 - .14 Using the 10-Fold method, 5 features are selected to obtain the best accuracy; using the LOO method, 9 features are selected to obtain the best accuracy; using 10-Fold method, 14 features are - .15 selected to obtain best ROC area; using LOO method, 13 features are selected to obtain best ROC area. - .16

### Development of prediction models

Eight prediction models were developed: KNN, SVM, LR and DNN models were all developed for both 7-feature and all-feature sets. The LR model with 7 features is shown in Table 5.

**Table 5.**
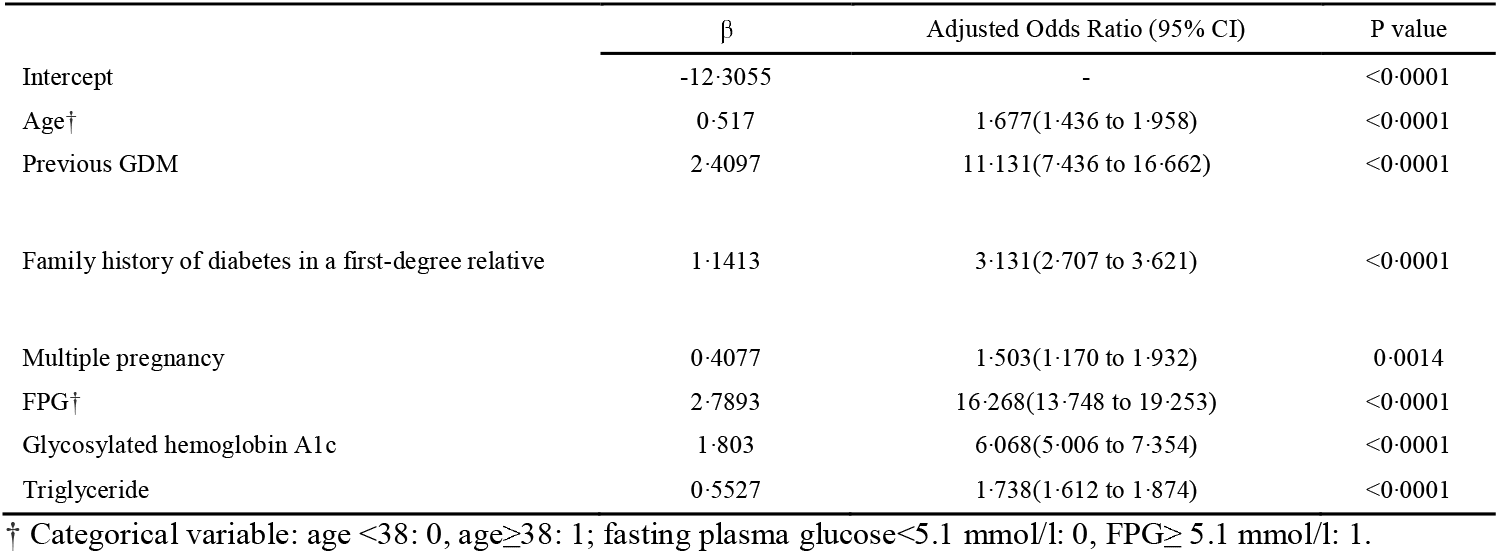
Multivariate analysis for the 7-feature LR model.

### Discrimination of different models

The AUC of different models is provided in Fig 3a and Table 6. All-feature DNN, SVM, KNN, and LR models obtained their best discriminative power with corresponding AUC of 0.92 (95%CI 0.91, 0.93), 0.82 (95%CI 0.81, 0.83), 0.63(95%CI 0.62, 0.64), and 0.85(95%CI 0.84, 0.85), respectively. The 7-feature DNN, SVM, KNN and LR models obtained their best discriminative power with corresponding AUC of 0.84(95%CI 0.83, 0.84), 0.69(95%CI 0.68, 0.70), 0.68 (95%CI 0.67, 0.69), 0.84(95% CI 0.83, 0.85).

**Table 6.** Sensitivity and specificity of different models.

| Prediction model | AUC[95%CI] | Best Pt threshold | Sensitivity | Specificity | Youden index |
| --- | --- | --- | --- | --- | --- |
| LR* | 0.84[0.83-0.85] | 0.17 | 71% | 80% | 0.51 |
| LR** | 0.85[0.84-0.86] | 0.02 | 67% | 86% | 0.53 |
| KNN* | 0.68[0.67-0.69] | - | 38% | 98% | 0.36 |
| KNN** | 0.63[0.62-0.64] | - | 28% | 98% | 0.26 |
| SVM* | 0.69[0.68-0.70] | 0.04 | 40% | 96% | 0.36 |
| SVM** | 0.82[0.81-0.83] | 0.05 | 61% | 89% | 0.5 |
| DNN* | 0.84[0.83-0.85] | 0.17 | 75% | 76% | 0.51 |
| DNN** | 0.92[0.91-0.93] | 0.2 | 94% | 81% | 0.75 |

\*: 7-feature model; \*\*: All-feature model

**Fig 3.**
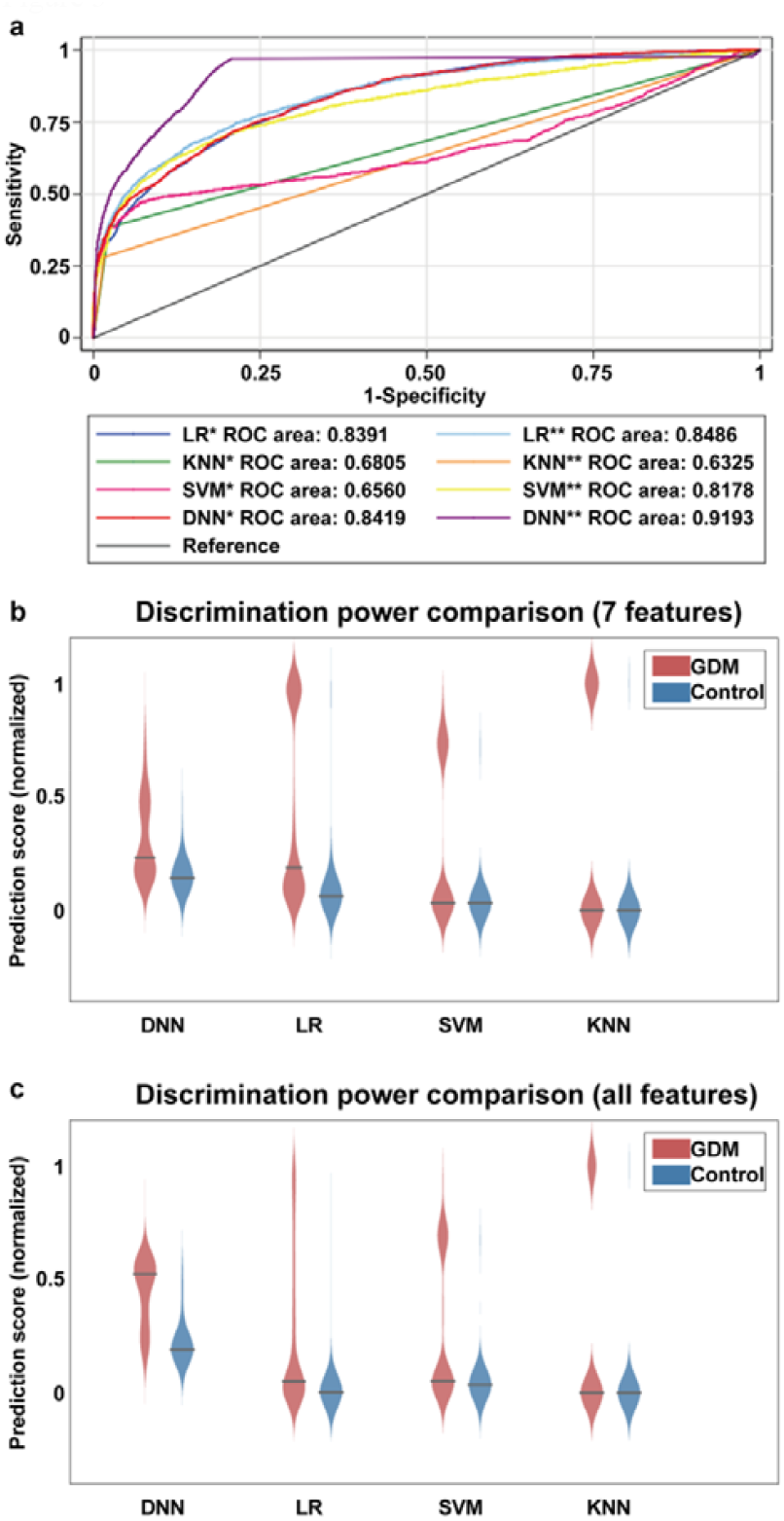
Discriminative power comparison between different prediction models. (a) The ROC curves of different prediction models based on the 7-feature panel (*) and all-feature panel (**). (b) and (c) are the violin plot comparisons of predicted score distribution using different prediction models, with 7-feature panel and all-feature panel, respectively.

The discrimination effect of each model is shown visually through violin plots (Figs 3b and 3c). The all-feature DNN demonstrated the best discrimination ability, and the traditional LR models resulted in higher AUC than KNN and SVM models. The optimal sensitivity and specificity of each model in certain Pt value ranges are given in Table 6. We also summarize previous prediction models or diagnostic criteria (Supplemental Table S1). Existing prediction models did not obtain the level of discrimination our LR and DNN models achieved (Supplemental Fig S1). In fact, the AUC of existing prediction models did not exceed 0.75.

### Calibration of different models

The HL test was used to test the calibration of the LR, SVM, and DNN models (Fig 4). KNN models could not provide risk probability for every case, so the HL test was not suited for KNN models. The P value of six different models was <0.0001 in HL test. The 7-feature models (Fig 4a, 4b, 4c) have better HL test performance than the all-feature models (Fig 4d, 4e, 4f). The 7-feature LR model revealed the most accurate calibration among all prediction models.

**Fig 4:**
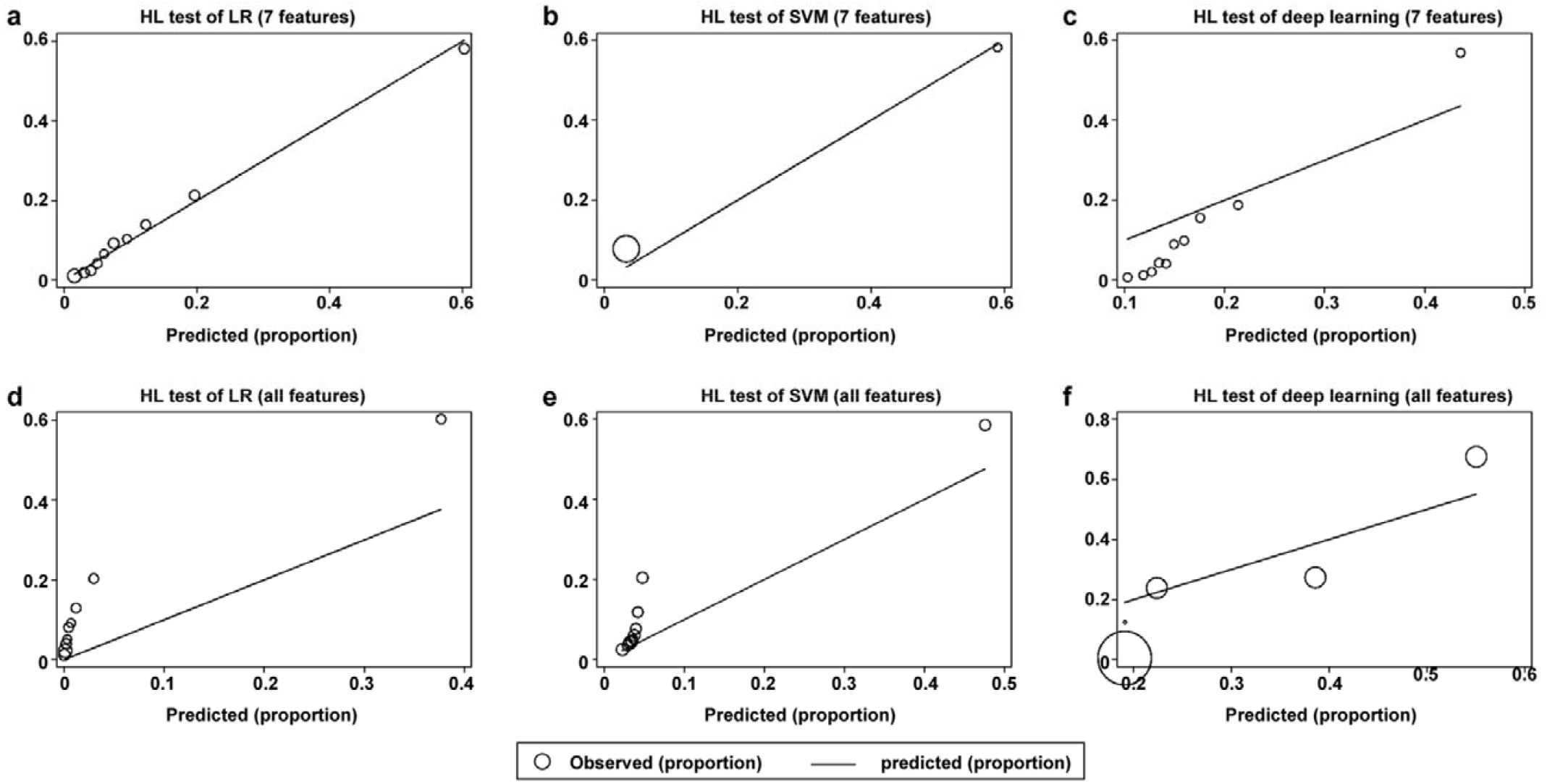
Calibration of different models. P value of all prediction models in HL tests is <0.0001. The 7-feature models (a, b, c) have better HL test performance than the versus all-feature models (d, e, f). Because if the model incorporates all features without selection, the model will inevitably over-fit, which will greatly reduce the calibration of models.

### Clinical use

The DCA of different models is presented in Fig 5. DCA is a useful method for evaluating the benefits of a prediction model across a range of patient preferences for accepting risks of overtreatment and undertreatment. This helps facilitate decisions about prediction, test selection, and use of these models. DCA showed that the range between the “treat-all” and “treat-none” line had a net benefit. For example, DCA showed that if the Pt value was set >4%, and <96%, using the 7-feature LR model to predict high risk status of GDM added more benefit than either the treat-all-patients scheme or the treat-none scheme.

**Fig 5:**
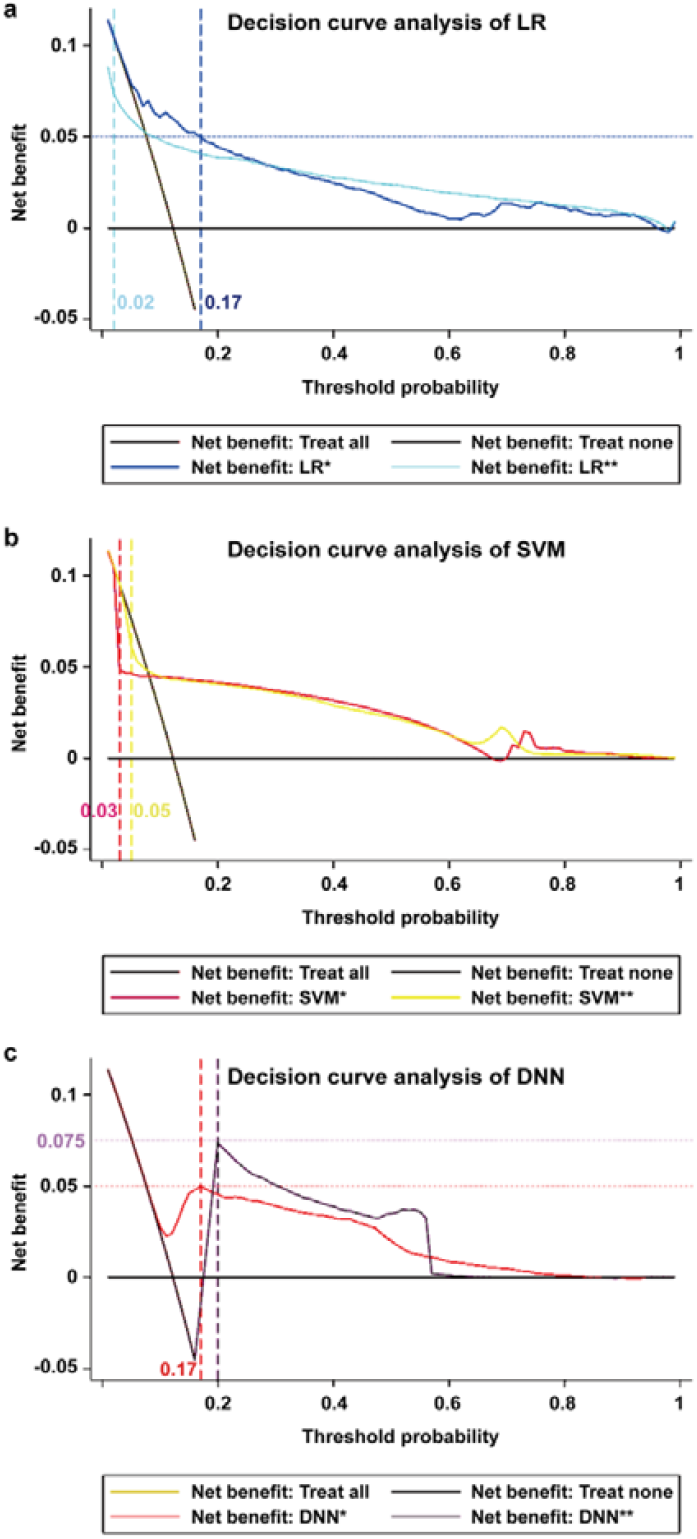
Decision curve analysis of different models. When prediction models have best discrimination (Pt sets at Youden index), 7-feature LR model and 7-feature DNN model can detect five positive GDM cases per 100 pregnant women in early pregnancy without increasing false positives (a, c), all-feature DNN model can detect 7.5 positive GDM cases per 100 pregnant women in early pregnancy without increasing false positive (c). While the net benefit brought by all-feature LR model and SVM models is no more than intervene-all-patients scheme under versus Pt value (a, b).

## Discussion

With the help of our model-free feature selection procedure, 17 features were screened out from the 73-feature set. Based on clinical experience and cost concerns, 7 features were finally chosen as the final predictive panel to construct AI models. Unlike many GDM prediction models previously used, this study added demographic data, blood biochemical indicators, and other data that can more easily be detected and collected during routine examinations. We validated previous predictive models in our validation group; the results further confirmed that our predictive model is more effective in terms of outcomes, such as AUC.

The 7-feature set includes age, personal history of GDM, family history of diabetes in a first-degree relative, fetal number, FPG, glycosylated hemoglobin A_1c,_ and triglyceride. Although the 7 features as a prediction panel has never been reported before, some of these features were individually reported as risk factors of GDM in previous researches [24, 25]. In addition to well-known risk factors, such as advanced age, previous GDM history, family history of diabetes, and blood glucose, our prediction model also included multiple pregnancy, HbA1c, and blood lipid levels. Twin pregnancies have an increased risk of GDM and higher rates of adverse pregnancy outcomes occur in GDM twin pregnancies [26]. HbA1c reflects blood glucose levels in the last 1-2 months, which circumvents the contingency brought by the single fasting blood glucose measurement [27, 28]. Previous studies have determined that insulin resistance is associated with obesity, the mechanisms involve dysregulated adipocytokines secreted by adipocytes that impair glucose tolerance and reduce insulin sensitivity [29, 30]. Adopting blood TG levels instead of BMI can better reflect the true obesity of pregnant women in early pregnancy, because BMI does not represent the body fat rate very well [31]. A meta-analysis shows that TG levels are significantly elevated among women with GDM compared to women without insulin resistance. This difference persists across all three trimesters of pregnancy [32], although the causal relationship between TG levels and GDM is still not clear [33]. The 7 features chosen in this study are not only significant from a computational point of view but are also supported by previous clinical studies. Interestingly, all 17 indicators selected by machine learning can be supported by relevant clinical literature, which also demonstrates that using machine learning to select features is an effective method.

Our study is the first GDM prediction model that combines feature selection, advanced machine learning technology, and traditional LR methods. Because our final prediction models were built upon a relatively small feature panel, the efficiency differences between prediction methods can be ignored. In terms of prediction efficacy, as shown in Fig 3 and Fig S1, DNN is better than LR, followed by SVM and KNN. The merit of DNN is that it can capture subtle nonlinear relationships between features and the outcome. However, the DNN approach may suffer from overfitting, and DNN is a black box to end-users, making it difficult to explain the contribution of each input features [34]. The LR model is more straightforward, where each feature’s contribution to the prediction is salient, especially in real time clinical practices where efficiency needs to be considered.

Our prediction model was designed to predict GDM based on the electronic health record of the first trimester, which is three months earlier than standard GDM diagnosis in 24-28^th^ week of pregnancy. The uncertain alteration of hormonal milieu during subsequent months increases the difficulty of predicting GDM via our LR model. The increased release of human placental growth hormone (hPGH), human chorionic somatomammotropin, progesterone, and placental growth hormone from placenta reprogrammed by maternal physiology to become insulin resistant in the second and third trimesters of pregnancy [35], are especially disruptive. Due to these limitations, it is impossible to predict all GDM patients in early pregnancy. However, our LR, SVM, and DNN models have a high success rate (AUC more than 0.80), and outperform existing early detection techniques.

The HL test was adopted to evaluate the calibration of prediction models [36]. The KNN model does not provide Pt value (it only provides “Yes” or “No”), so the HL test and the DCA curve did not contain KNN model result. The P values of all models for HL test were <0·0001, which implied that the model calibrations were not optimal. Excellent discrimination and dissatisfied calibration mean that the LR and DNN models are able to distinguish high risk status of GDM in early pregnancy, while the specific risk probability provided by these models can be improved further [37]. Although none of the models pass the HL test, the 7-feature LR model revealed better calibration than the DNN model. Notably, the deficiency of an appropriate assessment method for advanced machine learning models may lead to failure in calibration. Because the Pt values calculated from DNN and SVM models don’t represent risk probability, unlike the LR model, the HL test may be an inadequate method to evaluate the calibration of DNN and SVM models.

DCA was applied in this study for the assessment of clinical utility. This novel method offers insight for clinical consequences based on threshold probability (Pt) [38]. According to the results of the calculated DCA curves, the LR model and 7-feature DNN model were able to detect about 5 positive GDM cases per 100 pregnant women in early pregnancy without increasing false positives when the Pt value was set at 0.17 (Youden index). Generally, pregnant women with a high risk of GDM in the first trimester would receive physical activity and face-to-face nutrition intervention with nutritionists, rather than insulin injections [39,40]. The side effect of misdiagnosis brought by our models was slight, because lifestyle intervention is not an aggressive therapy. Therefore, relative low Pt values (0.17) are acceptable for doctors and patients. The all-feature DNN model revealed better DCA results when the Pt value is set to 0.2. Additionally, while the optimal Pt value (Youden index) was set at 0.03 for the 7-feature SVM model, 0.05 for the all-feature SVM model, and 0.02 for the all-feature LR model, these models added no additional benefit than the intervene-all-patients scheme.

Limitations of this study include a limited sample size and the fact that all the data used are collected from a single center. The diversity of laboratory testing between different hospitals caused by different laboratory instruments may also influence the effect of prediction and extrapolation. In future work, we will collect multi-center clinical data to verify the extrapolation of these prediction models. However, these shortcomings do not affect the demonstration that our feature selection and machine learning-based methodology is effective in early GDM prediction.

In conclusion, this study established prediction models in early pregnancy for the early intervention of GDM. Combining machine learning with clinical experience in feature selection has enormous utilization potential in establishing GDM prediction models based on demographic characteristics and laboratory testing. Our 7-feature LR and all-feature DNN prediction model have effective discrimination power for predicting GDM in early pregnancy. However, different modeling methods have distinct advantages and disadvantages, and the traditional LR model cannot be completely replaced yet.

## Data Availability

The data can be obtained from the corresponding author by email.

## Acknowledgments

We thank all the people who helped to collect the data and the graduate students who took part in the statistical analysis.

## Author contributions

Yan-Ting Wu contributed to the research idea and was the project coordinator. Chen-Jie Zhang contributed to the manuscript drafting. Ben Willem Mol contributed to manuscript modification. Cheng Li contributed to figures production and modification. Lei Chen contributed to the data export from the hospital electronic medical record system. Yu Wang, Jian-Zhong Sheng, and Jian-Xia Fan contributed to providing useful project suggestions. Yi Shi contributed to the machine learning algorithm and statistical analysis. He-Feng Huang contributed project guidance and financial support.

## Funding

This study was supported by the National Key Research and Development Program of China (2018YFC1002804, 2016YFC1000203), the National Natural Science Foundation of China (81671412, 81661128010), Foundation of Shanghai Municipal Commission of Health and Family Planning (20144Y0110), and Clinical Skills Improvement Foundation of Shanghai Jiaotong University School of Medicine (JQ201717).

## Ethic approval

This study was approved by the Medical Ethical Committee of International Peace Maternity and Child Health Hospital, School of Medicine, Shanghai Jiao Tong University (No. GKLW2019-05).

## Competing interests

No potential conflicts of interest relevant to this article were reported.

## Data availability

Please contact the corresponding author for the availability of the original data.

## Abbreviations

ADA: American Diabetes Association
AUC: Area under the curve
BMI: Body mass index
DCA: Decision curve analysis
DNN: Deep Neural Network
FPG: Fasting plasma glucose
GDM: Gestational diabetes mellitus
HbA_1c_: Glycosylated hemoglobin A1C
HDL: High density lipoprotein
HL: Hosmer-Lemeshow test
IADPSG: International Association of Diabetes and Pregnancy Study Groups
KNN: K-Nearest Neighboring
LR: Logistic Regression
SVM: Support Vector Machine
TG: Triglyceride

## Supplementary data

**Supplement table S1.** Risk factors, scoring system for the prediction of GDM and controversial diagnostic criteria reported in the literature.

| Study | Risk factors | Risk calculation/diagnostic criteria |
| --- | --- | --- |
| Naylor et al.(1), 1997 | Age | <30: 0; 31 ~ 34: 1; ≥35: 2 |
|  | BMI | <22: 0; 22.1 ~ 24.9: 1; ≥25: 2 |
|  | Race | White or black: 0; East Asian: 1; South Asian: 2 |
| Caliskan et al.(2), 2004 | Age | <25: 0; ≥25: 1 |
|  | BMI | <25: 0; ≥25: 1 |
|  | 1 st-degree relative with DM | No: 0; Yes:1 |
|  | Previous BW >4000g | No: 0; Yes:1 |
|  | History of adverse outcome | No: 0; Yes:1 |
| IADPSG(3), 2010 | Fasting plasma glucose in the early pregnancy | 5.1≤FPG<7.0mmol/L: GDM |
| van Leeuwen et al.(4), 2010 | BMI, race, previous GDM, 1 st- or 2 nd-degree relative with DM, | Probability of GDM = $1/[1+\exp(-b)]$; $b = (-6.1 + (0.13 \times \text{BMI}) + (0.5 \times \text{multipara with history of GDM}) - (0.67 \times \text{multipara without history of GDM}) + (0.83 \times \text{non-Caucasian ethnicity}))$ |
| Teede et al.(5), 2011 | Age | <25: 0; 25 ~ 34: 1; ≥35: 2 |
|  | BMI | <20: 0; 20 ~ 34.9: 1; ≥35: 2 |
|  | Race | White: 0; African, East Asian, and South Asian: 1 |
|  | 1 st-degree relative with DM | No: 0; Yes:1 |
|  | Previous GDM | No: 0; Yes:1 |
| Nanda et al.(6), 2011 | Age, BMI, race, previous GDM, previous BW >90th centile | Probability of GDM = $1/[1+\exp(-b)]$; $b = (-8.68947 + (0.10852 \times \text{BMI}) + (0.05365 \times \text{age}) + (1.00312 \times \text{South Asian}) + (0.88785 \times \text{East Asian}) + (3.72259 \times \text{previous GDM}) + (0.67673 \times \text{previous BW >90th centile}))$ |
| Sweeting et al.(7), 2017 | Age, BMI, race, previous GDM, parity, family history of DM | Probability of GDM = $1/[1+\exp(-b)]$; $b = (-5.9515 + (0.08462 \times \text{BMI}) + (0.0354 \times \text{age}) + (1.7033 \times \text{South Asian}) + (1.5500 \times \text{East Asian}) + (2.8624 \times \text{previous GDM}) + (3.0678 \times \text{family history of DM}) - (0.5836 \times \text{parity}))$ |

- 1.Naylor CD, Sermer M, Chen E, Farine D. Selective screening for gestational diabetes mellitus. Toronto Trihospital Gestational Diabetes Project Investigators. N Engl J Med 1997;337:1591–6. 2. Caliskan E, Kayikcioglu F, Ozturk N, Koc S, Haberal A. A population-based risk factor scoring will decrease unnecessary testing for the diagnosis of gestational diabetes mellitus. ActaObstetGynecol Scand. 2004;83:524–30. 3. Metzger BE, Gabbe SG, Persson B et al. International Association of Diabetes and Pregnancy Study Groups Consensus Panel. International Association of Diabetes and Pregnancy Study Groups recommendations on the diagnosis and classification of hyperglycemia in pregnancy. Diabetes Care. 2010;33:676–682. 4. van Leeuwen M, Opmeer BC, Zweers EJ, et al: Estimating the risk of gestational diabetes mellitus: a clinical prediction model based on patient characteristics and medical history. BJOG. 2010;117(1):69–75. - 5.Teede HJ, Harrison CL, Teh WT, Paul E, Allan CA. Gestational diabetes: development of an early risk prediction tool to facilitate opportunities for prevention. Aust N Z J ObstetGynaecol. 2011;51(6):499-504. - 6.Nanda S, Savvidou M, Syngelaki A, et al. Prediction of gestational diabetes mellitus by maternal factors and biomarkers at 11 to 13 weeks. PrenatDiagn.
7.Sweeting AN, Appelblom H, Ross GP, et al. First trimester prediction of gestational diabetes mellitus: A clinical model based on maternal demographic parameters. Diabetes Res ClinPract. 2017;127:44-50.

**Supplement Figure S1:**
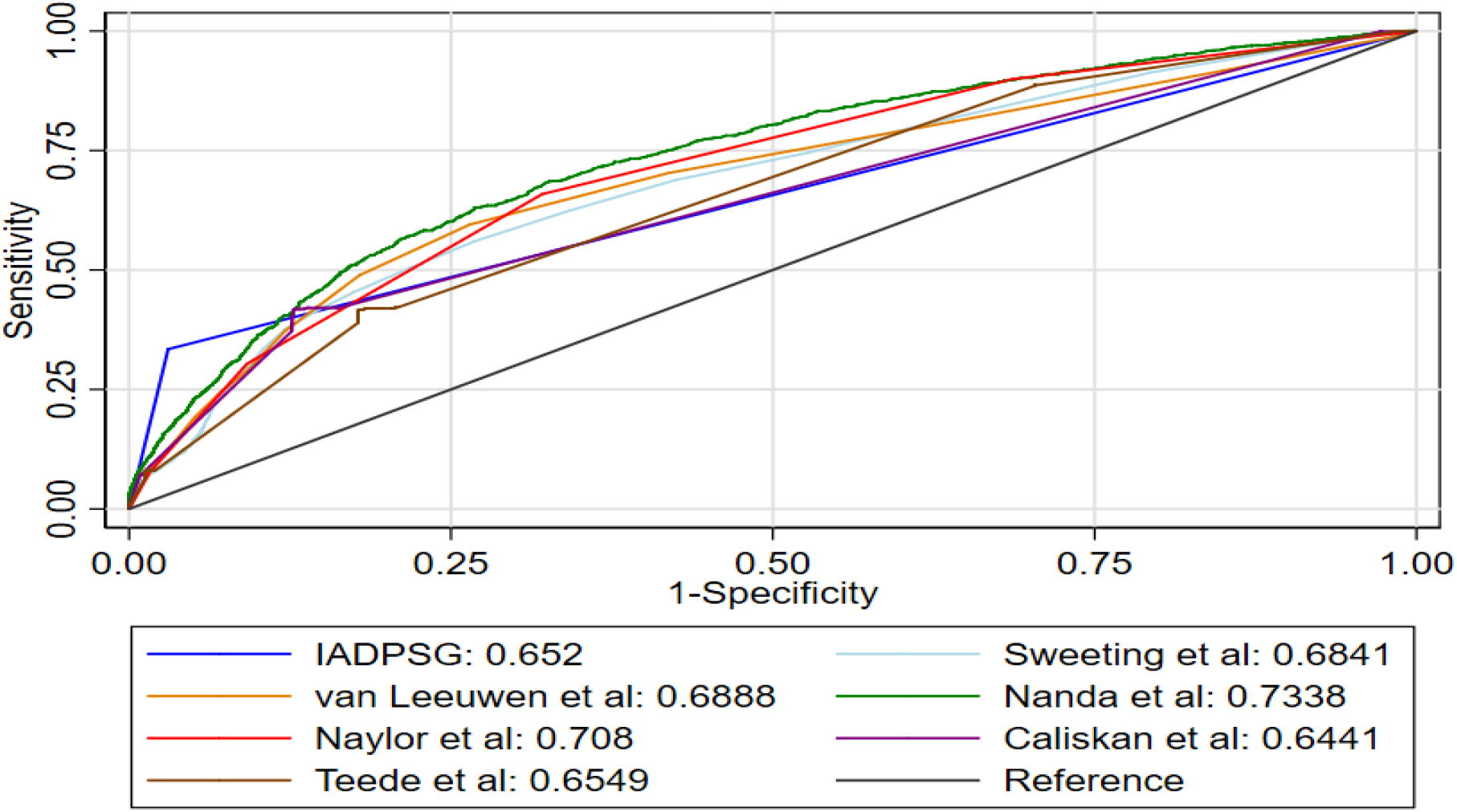
Discriminative power of previous models. The AUC of all previous prediction model or diagnostic criteria is less than 0.75. Nanda et al prediction model has the best AUC validated by our data, followed by Naylor et al model, van Leeuwen et al model, Sweeting et al model, IADPSG diagnostic criteria in the early pregnancy, Teede et al model, and Caliskan et al model.

## Notes

### Competing Interest Statement

The authors have declared no competing interest.

